# A Systematic Review on the Effect of Diabetes Mellitus on the Pharmacokinetics of Tuberculosis Drugs

**DOI:** 10.1101/2023.08.29.23294656

**Authors:** Muge Cevik, Ann Sturdy, Alberto Enrico Maraolo, Bart G.J. Dekkers, Onno W. Akkerman, Stephen H. Gillespie, Jan-Willem C. Alffenaar

## Abstract

**Objectives:** The coexistence of TB and DM (TB-DM) has been associated with an increased risk of treatment failure, death, delayed culture conversion and drug resistance. As plasma concentrations may influence clinical outcomes, we evaluated the evidence on the PK of TB drugs in DM individuals to guide management.

**Methods:** We performed a systematic review and meta-analysis through searches of major databases from 1946 to 6 July 2023. PROSPERO (CRD42022323566).

**Results:** Out of 4173 potentially relevant articles, we identified 16 studies assessing rifampicin PK, 9 on isoniazid, 8 on pyrazinamide and 3 on ethambutol. Two studies reported on second line anti-TB drugs. According to our analysis, RIF Tmax was significantly prolonged in patients with DM compared to non-DM patients. We found no significant differences for RIF C_max_, AUC _0-24_ or C2hr, INH C2hr, PZA C2hr, PZA T_max_ and ETB T_max_. While RIF C2hr was slightly reduced in TB-DM patients, this finding was not statistically significant.

**Conclusions:** This review comprehensively examines the impact of DM on the PK of TB drugs. We observed significant heterogeneity among studies. Given the association between lower plasma concentrations and poor clinical outcomes among DM patients, we recommend a higher dose limit to correct for larger bodyweight of patients with DM.

## INTRODUCTION

The global tuberculosis (TB) burden remains substantial, with more than 10 million people newly diagnosed per year. ^1^ Non-communicable diseases continue to pose challenges for both the management of TB and control strategies. For instance, over the next 10 years, the prevalence of Diabetes mellitus (DM) is estimated to double globally, with TB and DM co-epidemic surpassing the TB-HIV coinfection in low-middle income countries. ^2,3^ More alarmingly DM is increasing in the same population that is at high risk for developing TB.

DM is specifically important for the TB epidemic and future policies as DM has been shown to double the risk of acquiring TB ^4,5^ and DM patients have three-fold increased risk of progression to active disease, ^5^ highlighting that increased prevelance of DM will help to maintain the TB epidemic. In addition, TB-DM patients experience increased risk of severe disease, treatment failure, relapse and increased risk of death ^6,7^ as well as delayed sputum culture conversion and a greater risk of developing drug resistance. ^8,9^

There is however limited practical evidence to underpin treatment guidelines for TB-DM patients, such as length of treatment and drug dosing. This is important as plasma concentrations of TB medications may affect the clinical outcome. Zheng et al ^10^ showed an association between plasma concentrations of first line anti-TB drugs, culture conversion and clinical outcomes. Alfarisi et al. ^11^ demonstrated a positive association between time to culture conversion and isoniazid (INH) and rifampicin (RIF) concentrations and negative association with pyrazinamide (PZA) levels in DM patients. Pharmacokinetics (PK) of many other drugs are known to be altered in diabetic individuals. ^12^ DM may affect kidney function, gastric emptying and drug metabolism pathways. ^12^ This highlights the importance of understanding PK in TB-DM population. ^13^

In this systematic review and meta-analysis, we evaluated the available evidence on the PK of antituberculosis drugs in diabetic individuals to provide in-depth analysis to guide patient management.

## METHODS

### Search Strategy

We retrieved all original studies evaluating the effect of DM on the PK of all TB drugs recommended by the WHO for the treatment of drug-sensitive (DS) and drug-resistant TB. We systematically searched major databases including conference abstracts from 1946 to 6^th^ of July 2023 using Medical Subject Headings (MeSH) terms (Supplementary Material). We also manually screened the references of included original studies. To identify unpublished studies, http://clinicaltrials.gov was searched. This systematic review was conducted in accordance with the PRISMA statement. The protocol was registered at PROSPERO (CRD42022323566).

### Study Selection

Studies that reported PK of anti-TB medications among DM and non-DM participants were included. No restrictions on language and publication date were applied. General PK studies reporting a subgroup of DM participants were not eligible unless they included detailed PK parameters for DM and non-DM participants. Review articles, letters, case reports and case series with less than five participants were excluded, as were non-human studies.

### Data Extraction

Two authors screened and retrieved articles according to the eligibility criteria and performed full text review and final article selection. The following variables were extracted: demographics of comparator group(s) and the DM group including age, weight, and sex, study design, DM diagnosis, and DM medications, dose, dosing interval, sampling points, AUC, peak drug concentration (C_max_), half-life (t1/2), time to reach C_max_ (T_max_), volume of distribution (Vd) and clearance (CL) and were stratified by group. If these data were not reported, we also contacted the authors to request the data.

### Risk of bias in included studies

In the absence of available tools to assess risk of bias in PK studies, we assessed study quality and risk of bias using the ROBINS-I tool for non-randomised studies of interventions.^14^ Any disagreements regarding grading of quality were resolved through discussion with a third author.

### Analysis

Narrative synthesis of the findings was conducted and reported according to the Synthesis Without Meta-analysis (SWiM) guidelines. Descriptive statistics was used to describe key outcome measures. The pooled mean difference (MD) and 95% confidence interval (CI) was calculated for the main PK parameters of RIF, INH, PZA and ethambutol (ETB), through a random-effects model, when at least data from two studies were available. We compared values between TB patients with and without DM. Data expressed as medians (with interquartile range, IQR) were converted to meansl□and□standard deviations (SD).^15^ The pooling was performed according to an available case analysis scenario. Studies reporting means without SD values, medians without enough data to derive means and those reporting geometric means were excluded, since arithmetic means cannot be computed without raw data and log-transformed data cannot be mixed with untransformed ones in a meta-analysis.^16^ To express the amount of heterogeneity and estimate the between-study variance, when at least three studies were available for pooling a prediction interval (PI), was calculated.^17^ All calculations were carried out using R software, version 4.1.0, ‘meta’ package.

## RESULTS

4173 potentially relevant articles were identified. After removing duplicates, 3464 articles were retrieved for initial screening and 49 studies were included for full text review. After reviewing the eligibility criteria, 21 articles were included that assessed the PK of anti-TB medications among TB-DM patients. ^11,18-37^ Five included papers presented the same data: in conference abstract ^38-42^ and then in published form; ^19,25,29,32,39^ we included data from the published report. One study was only published as a conference abstract and the authors were not able to share data at the time of this review. ^43^ No relevant unpublished studies were found. The study selection process is recorded in a PRISMA flow diagram (**Figure 1**).

**Figure 1.**
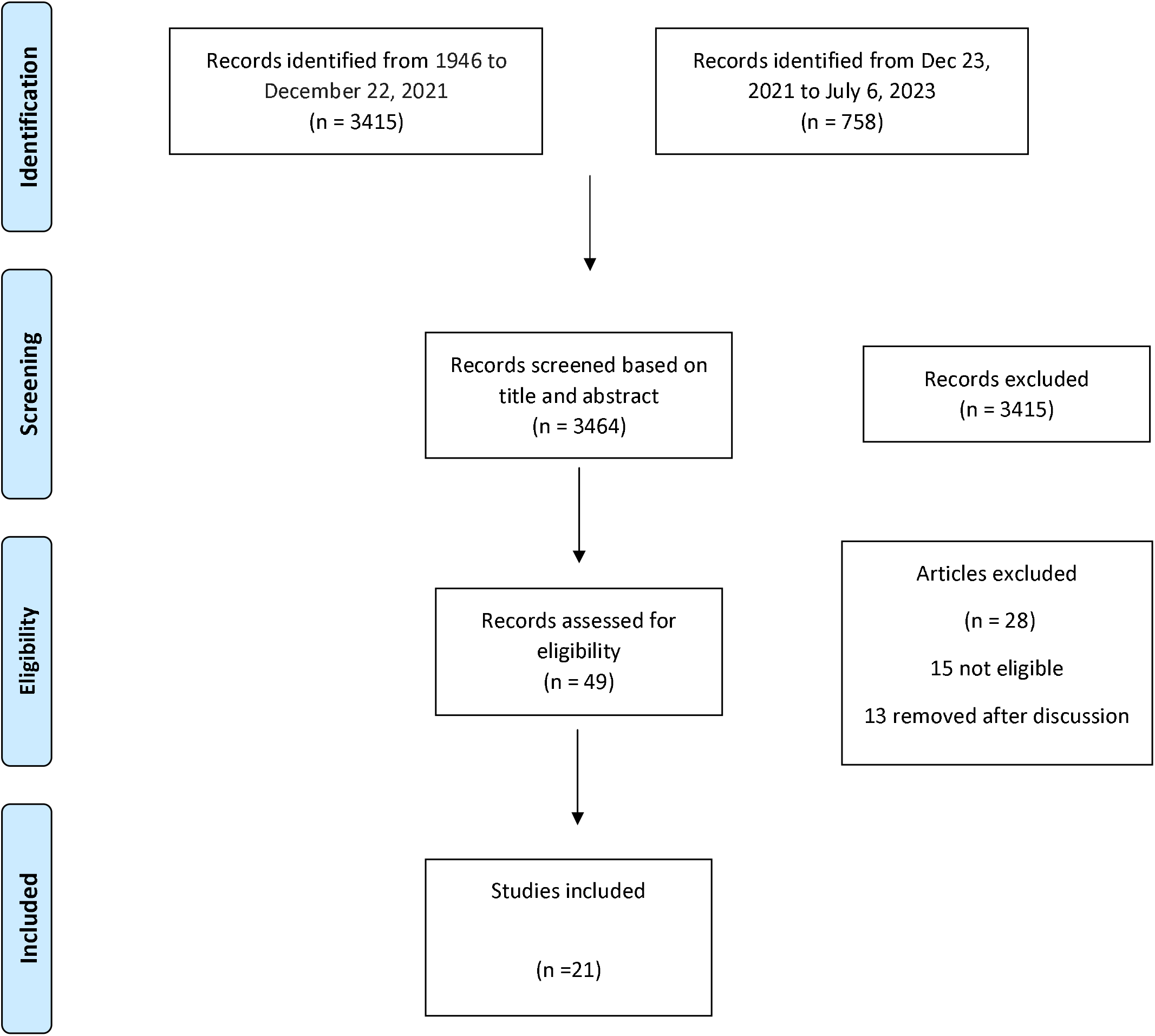
Flowchart describing study selection *From:* Moher D, Liberati A, Tetzlaff J, Altman DG, The PRISMA Group (2009). *P*referred *R*eporting *I*tems for *S*ystematic Reviews and *M*eta-*A*nalyses: The PRISMA Statement. PLoS Med 6(6): e1000097. doi:10.1371/journal.pmed1000097 For more information, visit www.prisma-statement.org.

### Risk of bias assessment

The risk of bias for each domain across all included studies is given in Supplementary material. While all studies have made a comparative analysis between DM and non-DM population, all had moderate to serious risk of bias in at least one domain. Overall, the existing body of literature examining PK of anti-TB medications among DM population is of medium quality.

### Rifampicin (Table 1)

Of the 21 studies, 16 measured RIF plasma concentrations, ^11,18,19,21,23,24,26-32,34-36^ majority of which were prospective PK studies except one retrospective study reporting results based on routinely collected data. ^24^

Out of 16 studies, 7 reported AUC values; three with intensive sampling (> 6 samples) ^27,28,34^ reporting AUC_0-24_ mg.h/L, three reported AUC_0-6_ mg.h/L, ^11,29,32^ and one reported AUC_0-8_ mg.h/L.^23^ Fifteen measured RIF peak concentrations (C_max_); mean absolute RIF C_max_ was below the recommended range (above 8 mg/L ^44^) in diabetic and non-diabetic groups in the majority of studies, and within the recommended range only in the 3 studies with intensive sampling. ^27,28,34^ Eight studies measured T_max_. In a study using population PK modelling, absorption rate constant (ka) was significantly increased and the volume of distribution (Vd) was significantly increased in the DM, ^19^ the authors inferring that the plasma concentrations would therefore be lower in the DM group. Another study found no difference in ka or Vd between the TB and TB-DM groups, but reduced clearance was observed and Vd was significantly higher in the TB-DM group when Vd was normalized to total body weight. ^27^

Raised blood glucose was inversely correlated with the rifampicin AUC/C_max_ in four studies, ^27,29,35,36^ whereas three other studies found no association.^21,28,34^ Two studies found that body weight and plasma concentrations were inversely correlated. ^18,29^

There were significant differences between DM and non-DM groups. Nine studies reported that DM patients had significantly higher BMI or weight ^11,18,21,23,26,27,29,32,35^ and nine studies reported the DM group to be older ^11,18,26-28,30,32,34,36^. One study that matched based on sex and age found C_max_ and AUC to be significantly lower and T_max_ significantly longer among DM group compared to non-DM. Eight studies reported DM patients to be on diabetes treatment ^11,18,21,26-30,36^, although no comparative analysis could be done as the majority of patients were receiving anti-diabetic medications.

Methodology differed between studies. Of the five studies finding a significantly decreased C_max_ in the diabetic group, four performed only a single 2-hour sampling ^18,24,35,36^, with one retrospective study ^24^ and one likely confounded by differences in weight (DM patients weighing more) ^29^.

DM patients had higher weight and/or BMI and some studies demonstrated a negative association between weight and plasma concentrations. The only study that matched based on weight (within 5 kg) ^34^ found no significant difference of RIF PK between groups. RIF doses used highly differed between studies which may have influenced the results, doses used detailed in Table 1.

### Isoniazid (Table 1)

Nine studies assessed isoniazid (INH) PK. ^11,18,22-24,26,28,31,33^ Of these, one was based on routinely collected retrospective data, ^24^ four studies used 600 mg dosing 3 times per week ^11,23,26,31^ and only 1 study sampled intensively. ^28^ Four studies assessed for NAT2 status using different methods. ^22,23,28,33^

Out of nine studies, four assessed the mean exposure; one AUC_0-24_ mg.h/L, ^28^ two AUC_0-6_ mg.h/L ^11,33^ and one AUC_0-8_ mg.h/L. ^23^ Four studies examined INH C_max_ ^11,18,26,28^ and three studies assessed T_max_ . ^23,33^

While one study reported concentrations to be negatively correlated with blood glucose, ^26^ in two studies INH C_max_/AUC was not correlated with FBG or HbA1c. ^22,28^ In one study INH concentrations were inversely correlated with weight. ^18^ There were significant differences between groups with the DM group being older and weighing more than non-DM group.

### Pyrazinamide (Table 1)

Pyrazinamide PK was reviewed in eight studies, ^11,18,23,25,26,28,31,34^ all of which were prospective PK studies. Patients in all studies except two were on 1500 mg PZA dosing, whilst one study used 1600 mg for patients >50 kg and 1200 mg for patients <50 kg ^28^ and other used 20-30 mg/kg. ^25^ Out of eight, four papers reported mean exposure to PZA; two reporting AUC_0-24_ mg.h/L, ^28,34^ one AUC_0-8_ mg.h/L ^23^ and one AUC_0-6_ mg.h/L. ^11^ Six studies reported C_max_ and three studies measured T_max_.

No correlation between FBG and AUC/C_max_ was observed in two studies ^28,34^ whereas one study showed a negative correlation.^26^ In a population PK modelling study, ^25^ increased apparent clearance was observed in TB-DM patients, most significantly in the group with patients >70 years and AUC_0-24_ mg.h/L was decreased in the DM group >70 years.

### Ethambutol (Table 1)

Three included studies assessed the effect of DM on EMB PK. ^18,28,34^ Two reported AUC_0-24_ mg.h/L and T_max_ ^28,34^, all reported C_max_.

### Other agents (Table 1)

Two studies assessed the effect of DM on cycloserine, ^37^ linezolid ^37^ and moxifloxacin. ^20^ A retrospective cross-sectional study reviewing the 2 h post-dose in routine practice found that 55% of the samples had below the lower limit of recommended cycloserine plasma concentrations and 17% had low linezolid concentrations. ^37^ DM patients had a lower cycloserine exposure, although this was not statistically significant, and there was no association between linezolid exposure and DM. In a recent retrospective study evaluating moxifloxacin PK, AUC_0–24h_ was shown to be significantly lower in patients with DM compared to age, sex and RIF matched TB patients without DM. ^20^ In line, peak and trough concentrations were also reduced in DM patients. Although the drug absorption, volume of distribution and T_max_ were comparable between TB-DM and TB patients, moxifloxacin clearance was increased in TB-DM patients.

### Meta-analysis

A quantitative pooling was possible in seven cases, of which four involved RIF. We pooled data from four studies reporting RIF T_max_.^27-30,34^ Overall, RIF T_max_ was significantly higher in the TB-DM group: MD 0.65 (95% CI, 0.03-1.27) (Figure 2). Two studies were pooled for RIF AUC_0-24_,^27,30^ four for RIF C_max_ ^21,23,27,30^ and four for RIF C_2hr_ ^18,26,31,35^, and no statistically significant differences were detected (Figure 2-3). However, RIF C_2hr_ was lower in TB-DM group with a MD of -1.68 (95% CI, -5.41; 2.05). In regard to other first line drugs, we observed no significant differences for INH C2hr, PZA C2hr, PZA T_max_ and ETB T_max_ (supplementary material).

**Figure 2:** Meta-analysis of Rifampicin (RIF AUC0-24 and RIF Tmax)

**Figure 3:** Meta-analysis of Rifampicin (RIF Cmax and C2hr)

## DISCUSSION

This systematic review provides a comprehensive account of the impact of DM on the PK of anti-TB medications. While the evidence is heterogeneious, according to our analysis, RIF T_max_ is significantly prolonged in patients with DM compared to non-DM patients; however, PI crossed the no-effect threshold, indicating that in future studies opposite results may be determined. Whereas we found no significant differences between DM and non-DM patients for RIF C_max_, AUC _0-24_ or C2hr, INH C2hr, PZA C2hr, PZA T_max_ and ETB T_max_. While RIF C2hr was slightly reduced in TB-DM patients, this finding was not statistically significant.

Diabetes predominantly effects gastric motility via neuropathy.^12^ This would be postulated to prolong T_max_ rather than reduce the overall AUC. Our findings of prolonged RIF T_max_ may be a reflection of the impact of DM on gastric motility and delay in drug absorption. However, the efficacy of most of the anti-TB medications is driven by AUC, which is difficult to determine with 1 or 2 sampling timepoints. Therefore, interpretation of studies with limited sampling points should be with caution. Studies with more than one sampling point, ideally with intensive sampling comparing DM vs non-DM population, or limited sampling strategy using population PK modelling will guide our management strategy in this population.

There is some evidence that PK among DM patients, may have clinical relevance in terms of MIC and culture conversion. In a retrospective study, where all DM patients underwent TDM and dose adjustment accordingly, time to culture conversion improved significantly (average 19 days earlier) among DM patients. ^45^ Recently, studies with higher doses of RIF has demonstrated bacteriologically sustained cure in a general TB population.^46^ Perhaps these results and the higher RIF strategy have more relevance for the TB-DM population in which we observe more heterogenous plasma concentrations and that reducing the bacterial burden quicker may result in reduced relapse rates.

We observed high variability in findings and significant heterogeneity among studies. Firstly, there were differences in terms of demographics; age, sex and body weight, DM diagnosis and anti-DM medications. Increased body surface area distribution, delayed drug absorption due to DM related gastroparesis and changes in expression of enzymes involved in metabolism ^47^ are major factors that may contribute to altered PK. ^12,48^ In most studies, DM patients weighted more than non-DM controls. However, matching weight is unlikely to eliminate potential biases as DM participants continued to receive capped dosing despite weight differences. Screenning DM at the time of TB diagnosis may result in some with transient hyperglycaemia being identified as DM. While most of the patients were on anti-diabetic medications, the degree of diabetic control was not provided, limiting our understanding.

Secondly, PK sampling methodology and drug doses used varied widely across studies. Those with a single 2 hr sampling point assume T_max_ to occur at 2 hrs (C_max_). Therefore, studies without intensive sampling may not accurately capture the C_max_. However, it is important to note T_max_ is highly variable for RIF in general. Particularly affecting the isoniazid studies, dosing differed markedly, with 4 out of 9 using 600 mg 3 times/week, which is no longer recommended. Only four out of 9 studies made any assessment of NAT2 acetylator status which is a key variable in isoniazid PK likely to confound results. ^49^

We identified another systematic review assessing the impact of DM on the PK of rifampicin among TB patients ^50^. The review included studies published up until September 2020 and identified seven studies from which pooled estimates were calculated. The same authors updated their systematic review including studies with C_max_ at 2hr, ^51^ and identified 17 studies reporting RIF plasma concentrations.

Our study has limitations. Firstly, we identified a limited number of PK studies despite a comprehensive literature search. Secondly, due to studies reporting geometric means we were unable to pool data from those reporting AUC 0-24. Finally, we did not include general PK studies reporting on TB-DM patients as a subgroup.

Given the association observed between dose adjustment and clinical improvement, dose adjustment based on TDM may be beneficial in this patient population. We recognise that in most global settings TDM is not easily available. Until more simplified and cost-effective tests are easily available, ^52^ the alternative would be to empirically increase drug doses in all individuals with DM. While TB drug exposures in diabetes is mixed, given RIF has a wide therapeutic window ^53^ and successful treatment outcome depends on sufficient drug exposure, we recommend considering a higher dose limit for patients with DM; instead of capped dosing, using 10mg/kg to be able to correct for larger bodyweight of patients with DM. This is also a group that should be prioritised in ongoing studies addressing empirically increasing doses of TB medications. In order to achieve definitive answers for PK in DM population, we recommend important considerations as in **Table 2**.

## Supporting information

supplementary materiaal

## Data Availability

All data produced in the present study are available upon reasonable request to the authors

## Acknowledgements

This work was supported by the Chief Scientist Office (CAF/20/03) and British Infection Association (Grant/2022/SPG/MC) received by MC.

## Author contributions

MC conceptualized the scope of the review, led the systematic review, literature search, data collection and wrote the first and subsequent drafts of the manuscript. AS involved in creating the table from selected papers. AEM performed the meta-analysis on selected papers. All authors contributed to the final version of the manuscript and approved it for publication.

## Figures and tables

Table 1: Summary of included studies

Table 2: Recommendations for future research

