## supplementary material for "A Systematic Review on the Effect of Diabetes Mellitus on the Pharmacokinetics of Tuberculosis Drugs"

Search strategy

The following databases will be searched: MEDLINE, EMBASE, Web of Science and Scopus. For the following conferences we will hand search abstracts from: European Congress of Clinical Microbiology and Infectious Diseases (ECCMID). Additionally, if any literature reviews are identified, reference lists of those review articles will be searched.

 Search strategy :

1            exp Diabetes Mellitus/

2            diabetes.mp.

3            exp Tuberculosis/ or tb.mp.

4            tuberculo*.mp.

5            1 or 2

6            3 or 4

7            5 and 6

8            drug concentration*.mp.

9            exp Pharmacokinetics/

10          exp Drug Monitoring/

11          pharma*.mp

12          8 or 9 or 10 or 11

13          7 and 12

Restrictions:

- There will be no restriction by language or time.
