## supplementary material for "A Systematic Review on the Effect of Diabetes Mellitus on the Pharmacokinetics of Tuberculosis Drugs"

### Effect of diabetes mellitus on the pharmacokinetics of first-line tuberculosis drugs

#### Citation

Muge Cevik, Bart Dekkers, OW Akkerman, Johannes Alffenaar, Stephen Gillespie. Effect of diabetes mellitus on the pharmacokinetics of first-line tuberculosis drugs. PROSPERO 2022 CRD42022323566 Available from: https://www.crd.york.ac.uk/prospero/display_record.php?ID=CRD42022323566

#### Review question

Primary question: what is the effect of diabetes mellitus (DM) on the pharmacokinetics of first-line tuberculosis (TB) drugs?

#### Searches

The following databases will be searched: MEDLINE, EMBASE, Web of Science and Scopus.

For the following conferences we will hand search abstracts from: European Congress of Clinical Microbiology and Infectious Diseases (ECCMID).

Also, if any literature reviews are identified, the reference lists of those review articles will be searched.

We have no restrictions on the type of study included however envisage that the majority of literature will be case reports, case series and possibly cohort studies. There are no exclusion criteria in terms of study design.

Additional search strategy information can be found in the attached PDF document (link provided below).

#### Types of study to be included

We have no restrictions on the type of study included however envisage that the majority of literature will be case reports, case series and possibly cohort studies. There are no exclusion criteria in terms of study design.

#### Condition or domain being studied

Tuberculosis and diabetes mellitus.

#### Participants/population

Inclusion criteria:

- Studies that report on pharmacokinetics of first line treatment in TB patients with diabetes mellitus;

- The review participants will be tuberculosis and diabetic patients of all ages and genders.

There are no limitations such as age and gender.

#### Intervention(s), exposure(s)

Exposure: the various categories of diabetes mellitus including type 1 diabetes, type 2 diabetes, gestational diabetes mellitus, and other types of diabetes.

#### Comparator(s)/control

Individuals infected with TB without diabetes.

#### Context

No limitations on study settings will be made, including inpatient and outpatient settings in any country globally.

#### Main outcome(s)

The main outcome is reporting the pharmacokinetics of first line tuberculosis drugs among patients with diabetes mellitus to better understand the influence of DM on the TB treatment outcomes.

We do not anticipate that there will be sufficient data to permit meta-analysis. Any quantitative data analysis completed will be limited to descriptive statistics.

#### Measures of effect

Not applicable.

#### Additional outcome(s)

None.

#### Data extraction (selection and coding)

Two authors will independently review reports by title and abstract for relevance, with at least 20% of all reports being screened in duplicate to ensure continuity. The full text reports of studies not excluded by title and abstract will then be reviewed by two authors independently to consider if they are eligible to be included. Any disagreements regarding study inclusion will be resolved through discussion with a third author.

Following this, data will be extracted used a standardised electronic data extraction form. We will extract and record the following information, where available, for each included study:

1. Bibliographic: first author, year;

2. Methods: study design, setting, country;

3. Participants: number, age, gender, sexuality (if reported), co-morbidities, drugs (e.g. steroids);

4. Diabetes: screening;

5. Tuberculosis: treatment regimen used, treatment doses;

6. Pharmacokinetics: number of observation points, key observations for PK parameters (AUC0-24h, Cmax, t½, clearance, and volume of distribution).

#### Risk of bias (quality) assessment

We have attempted to reduce biases in the search, such as reducing reporting bias through inclusion of grey literature.

We acknowledge that limiting the search to reports in English may introduce some bias to our results.

From preliminary searches we anticipate that most included studies will be case-control studies or cross-sectional studies and the risk of bias will be assessed using the Joanna Briggs Institute Manual for Evidence Synthesis. If cohort studies and/or randomised controlled trials are included, then risk of bias will be formally assessed using the GRADE tool.

#### Strategy for data synthesis

The studies will be summarised in the text and table form, a synthesis of the findings will be conducted and reported according to the Synthesis Without Meta-analysis (SWiM) guidelines (Campbell et al 2019). The syntheses will be structured according to study design (if studies other than case reports and case series are included), type of coronavirus, viral shedding parameters, and type of coronavirus outcome measures reported.

Descriptive statistics will be used to describe key outcome measures, AUC0-24h, Cmax, t½, clearance, and volume of distribution of tuberculosis drugs in TB patients with DM compared to those without DM. It is anticipated that the data will be too heterogenous to permit further quantitative synthesis in a meaningful way.

Provided two or more studies are identified, then the data synthesis will be undertaken, although if the number of included studies is very small then the data available for synthesis in for each outcome measure will be very limited.

A narrative synthesis of the findings will be conducted. The characteristics of patients in the included studies will be presented in a table.

#### Analysis of subgroups or subsets

None planned.

#### Contact details for further information

Muge Cevik

#### Organisational affiliation of the review

University of St Andrews

#### Review team members and their organisational affiliations

Dr Muge Cevik. University of St Andrews

Dr Bart Dekkers. University of Groningen

Professor OW Akkerman. University of Groningen

Professor Johannes Alffenaar. University of Sydney

Professor Stephen Gillespie. University of St Andrews

#### Type and method of review

Intervention, Narrative synthesis, Systematic review

#### Anticipated or actual start date

21 December 2021

#### Anticipated completion date

30 May 2022

#### Funding sources/sponsors

No funding sources for this review

#### Conflicts of interest

#### Language

English

#### Country

Australia, Netherlands, Scotland

#### Stage of review

Review Ongoing

#### Subject index terms status

Subject indexing assigned by CRD

#### Subject index terms

Diabetes, Gestational; Diabetes Mellitus, Type 1; Diabetes Mellitus, Type 2; Drug Therapy; Humans; Pharmacokinetics; Treatment Outcome; Tuberculosis

#### Date of registration in PROSPERO

06 April 2022

#### Date of first submission

06 April 2022

#### Stage of review at time of this submission

| Stage | Started | Completed |
|---|---|---|
| Preliminary searches | Yes | No |
| Piloting of the study selection process | Yes | No |
| Formal screening of search results against eligibility criteria | Yes | No |
| Data extraction | No | No |
| Risk of bias (quality) assessment | No | No |
| Data analysis | No | No |

*The record owner confirms that the information they have supplied for this submission is accurate and complete and they understand that deliberate provision of inaccurate information or omission of data may be construed as scientific misconduct.*

*The record owner confirms that they will update the status of the review when it is completed and will add publication details in due course.*

#### Versions

06 April 2022
